# Early metabolic disruption and predictive biomarkers of delayed-cerebral ischemia in aneurysmal subarachnoid haemorrhage

**DOI:** 10.1101/2023.04.26.23289184

**Authors:** Karim Chikh, David Tonon, Thibaut Triglia, David Lagier, Anouk Buisson, Marie-Christine Alessi, Catherine Defoort, Sherazade Benatia, Lionel J Velly, Nicolas Bruder, Jean-Charles Martin

## Abstract

**BACKGROUND:** Delayed cerebral ischaemia (DCI) following aneurysmal subarachnoid haemorrhage (aSAH) is a major cause of complications and death. Here we set out to identify high-performance predictive biomarkers of DCI and its underlying metabolic disruptions using metabolomics and lipidomics approaches.

**METHODS:** This single-centre retrospective observational study enrolled 61 consecutive patients with severe aSAH requiring external ventricular drainage between 2013 and 2016. Of these 61 patients, 22 experienced a DCI and were classified as DCI+ and the other 39 patients were classified as DCI-. A further 9 patients with other neurological features were included as non aSAH controls. Blood and cerebrospinal fluid (CSF) were sampled within the first 24 h after admission. We carried out LC-MS/MS-based plasma and CSF metabolomic profiling together with total lipid fatty acids analysis.

**RESULTS:** We identified a panel of 20 metabolites that together showed high predictive performance for DCI (area under the receiver operating characteristic curve: 0.968, specificity: 0.88, sensitivity: 0.94). This panel of metabolites included lactate, cotinine, salicylate, 6 phosphatidylcholines, and 4 sphingomyelins. Analysis of the whole set of metabolites to highlight early biological disruptions that might explain the subsequent DCI found peripheral hypoxia driven mainly by higher blood lactate, arginine and proline metabolism likely associated to vascular NO, dysregulation of the citric acid cycle in the brain, defective peripheral energy metabolism and disrupted ceramide/sphingolipid metabolism. We also unexpectedly found a potential influence of gut microbiota on the onset of DCI.

**CONCLUSION:** We identified a high-performance predictive metabolomic/lipidomic signature of further DCI in aSAH patients at admission to a NeuroCritical Care Unit. This signature is associated with significant peripheral and cerebral biological dysregulations. We also found evidence, for the first time, pointing to a possible gut microbiota/brain DCI axis, and proposed the putative microorganisms involved.

**Clinical trial registration:** Clinicaltrials.gov identifier: NCT02397759

## Introduction

Delayed cerebral ischaemia (DCI) occurs in about 30% of patients with aneurysmal subarachnoid haemorrhage (aSAH) within the first 2 weeks after the haemorrhage (1). DCI is responsible for increased morbidity and mortality (2). It is crucial to identify patients at risk of developing DCI after aSAH in order to develop targeted therapies for treating or preventing this potentially life-threatning complication. However, there is still no established biomarker for predicting DCI. Furthermore, there is a broader need to more clearly establish the early biological disruptions that ultimately lead to DCI, as only hypotheses have so far been put forward. DCI was long thought to be caused by cerebral vasospasm, but recent studies support the notion it is a multifactorial pathophysiology that includes cerebral vascular dysregulation, microthrombosis, cortical spreading depolarisation, and neuroinflammation (3).

Metabolomics and its subfield lipidomics are powerful tools for identifying putative biomarkers in various different contexts, from disease diagnostics (4) and disease risk analysis (5,6) to prediction of therapeutic response (7). Moreover, changes in metabolome provide a molecular snapshot of cellular activity and thus provide important clues to understanding functional changes in the metabolic pathways that drive disease risk.

Modifications of some metabolites and lipid levels have been reported in the context of DCI or vasospasm associated with aSAH. Elevated CSF levels of ceramides (8), arachidonic acid, linoleic acid and palmitic acid (9), elevated lactate/pyruvate ratio (10) and taurine in cerebral microdialysis samples (11), high blood lactate levels and glucose levels (12), and elevated plasma taurine levels (13) have all been reported in aSAH patients who later develop cerebral vasospasm and/or DCI. Nevertheless, there is no consensual biomarkers accepted for routine use in clinical practice (14).

In a previous paper, we reported an early increase in CSF MMP-9 concentrations in patients who later developed DCI (15). Here we carried out both LC-MS/MS-based plasma and CSF metabolomic profiling and gas-chromatography total lipid fatty acids analysis in our cohort of patients presenting aSAH with and without DCI. Our objectives were to identify predictive biomarkers of DCI and to decipher the early underlying metabolic pathway disruptions leading to DCI.

## Methods

### The study complies to the STROBE and Machine Learning Predictive Models guidelines Study population and retrospective study setting

After obtaining written informed consent from next of kin, we enrolled 61 consecutive patients with spontaneous aSAH who met the inclusion criteria. To be eligible for inclusion, patients had to be over 18 years old and admitted to the NCCU of La Timone university hospital (Marseille) within 24h post-bleeding, and their conditions had to require clinically indicated external ventricular drainage. A full description of the inclusion and exclusion criteria, data collected, radiographic characteristics, criteria used for cerebral vasospasm and DCI determination can be found in supplemental methods. A further 9 patients without aSAH admitted to the NCCU for other neurological disorders (**see Table S1**) were included as controls (n=9). Blood and CSF were sampled in the first 24 h post-aSAH. CSF samples were collected from the external ventricular drain into anticoagulant-free sterile tubes. Blood samples were collected in sterile citrate tubes. Samples were centrifuged for 10min at 2500g at 4°C then immediately transferred to cryotubes and stored at −80°C until analysis.

### Sample preparation

For metabolomics analysis, 100 µL of CSF or plasma samples was protein-precipitated with 400 µL cold methanol (−20°C). For lipidomics analysis, compounds were extracted using the conventional Folch extraction method by adding 800 μL of ice-cold chloroform:methanol (1:1 v/v) to 100 μL of plasma or CSF (see supplemental methods for details).

### LC–MS conditions and informatics processing for metabolomics and lipidomics analysis

LC–MS analyses were performed on a Dionex Ultimate 3000 (Thermo Fisher Scientific, San Jose, CA) ultra-performance liquid chromatography (UPLC) system coupled with a Thermo Q Exactive Plus mass spectrometer (MS) (see supplemental methods for details). After LC–MS acquisition, data was processed using the R package XCMS (https://bioconductor.org/packages/release/bioc/html/xcms.html). Polar metabolites were annotated using an internal database and lipids were identified using LipidSearch 4.1 software (Thermo Fisher Scientific, San Jose, CA).

### Total lipid fatty acid quantification

Total lipid plasma and CSF fatty acid composition were analysed using a gas chromatography system with flame ionisation detection. The method is described in full in supplemental methods.

### Statistical analysis

Full details of all the statistical methods used can be found in supplemental methods. Statistical analyses, univariate and multivariate regressions, hierarchical PLS-DA modelling, and pathway enrichment analysis were performed using MetaboAnalyst 5.0 (http://www.metaboanalyst.ca), SIMCA P12 (Umetrics, Umea, Sweden) and LION (http://www.lipidontology.com). Venn plots were constructed using the Venny 2.0.2 online tool (bioinfogp.cnp.csic.es/tools/venny). Partial correlations were calculated with the R package GeneNet, and network visualisation was performed using Cytoscape. The biomarker identification process only used data from 50 patients, as CSF plus plasma data was not available for 11 patients. A DCI+ score equation to predict the clinical status of each patient was calculating using the partial least squares (PLS) algorithm combining all the individual discriminating variables, as described elsewhere (5,16).

## Results

### Clinical features of the studied population

Among all the patients with aSAH admitted to our NCCU between 2013 and 2016, 61 patients met the inclusion criteria. Of these 61 patients, 22 developed DCI (DCI+ group) whereas 39 did not (DCI-group). Patient-sample characteristics are reported in supplemental Table 2 and supplemental Figure S1. The only significant between-group difference in demographic variables was for patient age: DCI+ patients were significantly younger than DCI-patients. The DCI+ group tended to have more smokers and more cases with migraine. Vasospasm was identified in 90.1% of DCI+ patients but none in DCI-patients (p<0.001).

### Feature detection in metabolomics and lipidomics analysis and fatty acid content

A total of 180 and 153 annotated metabolites and 166 and 290 annotated lipids were identified in CSF and plasma, respectively. In addition to metabolomics and lipidomics, 27 fatty acids were quantified in CSF and plasma by a targeted method. A unique data matrix featuring 843 variables was built with all these compounds and used for further statistical analyses. Missing values (4 in plasma and 16 in CSF) for fatty acids were replaced by an estimate based on Bayesian principal component analysis.

### Biomarker selection and validation

As both CSF and plasma were not available for 6 and 5 patients, respectively, the biomarker identification process only used data from 50 patients of our cohort (33 DCI- and 17 DCI+). To select and validate a set of biomarkers, we applied an iterative workflow with randomly left-out patients in multiple predictive PLS-DA models (5,16). PLS regression also allowed to predict class assignments for unknown patients. A list of 20 metabolites was selected based on the most commonly shared variables found in 8 consecutively constructed PLS-DA training models in which four DCI-patients and two DCI+ patients were repeatedly and randomly left out. In each model, the variables were selected based on the shift in the PLS variable importance in projection score (VIP) from the normal distribution. The accuracy, goodness-of-fit R2 and goodness-of-prediction Q2 of the final PLS-DA model comprising all the individuals and based on these 20 selected metabolites were 0.89, 0.69 and 0.55, respectively (Figure 1A). All these 20 metabolites were significantly different between DCI- and DCI+ on univariate *t*-test (FDR adjusted *p*-values < 0.05): 12 of these metabolites were relatively higher in abundance and 8 were relatively lower in abundance in DCI+ patients, and only 3 of them came from CSF (Figure 1B).

**Figure 1.**
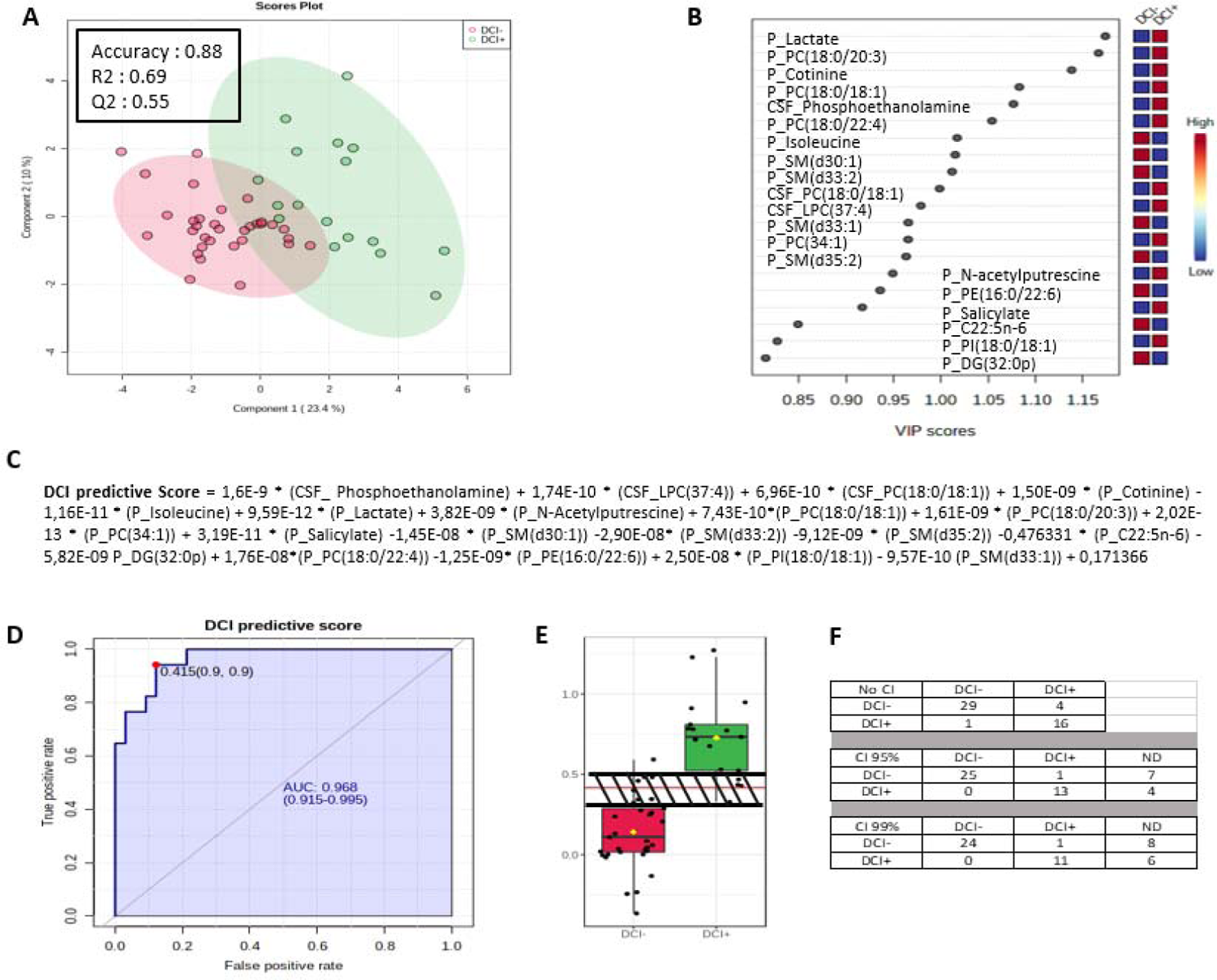
DCI predictive score and biomarker analysis. **A**: PLS-DA 2D scores plot showing distribution of DCI+ and DCI-patients according to principal components 1 and 2. **B**: VIP scores plot of the 20 metabolites selected in the final PLS-DA model. P and CSF prefixes for metabolite names indicate that the corresponding metabolites were detected in plasma and cerebrospinal fluid, respectively. **C:** Equation of the composite score for prediction of DCI status. P and CSF prefixes indicates if the corresponding metabolites are detected respectively in plasma or CSF. **D:** Receiver operating characteristic curve associated with DCI predictive score. The red point represents the best cut-off according to sensitivity and specificity. AUC: area under the curve. **E:** DCI predictive scores plot. The red line represents the test cut-off (0.415). The shaded area represents the 95% confidence interval (CI) grey zone. **F:** DCI status prediction according to DCI predictive score when applying no CI, 95% CI, and 99% CI. ND: not determined. Number of patients in each observed DCI status group is given in the corresponding rows. Number of patients in each predicted DCI status group is given in corresponding columns.

The selected metabolites were subsequently combined to generate a meaningful clinical composite score for each individual. This predictive score was calculated from the PLS algorithm using the PLS partial correlation coefficients applied to each metabolite, with DCI status used as the predicted variable (Figure 1C). Working up from this equation, we calculated a predictive DCI score for each individual and tested each score using a receiver operating characteristic curve (Figure 1D). Area under the curve was 0.968, specificity was 0.88, sensitivity was 0.94, and the cut-off threshold score above which DCI+ individuals distinguished from DCI-ones was 0.415 (Figure 1D). When using the strict cut off value, 86% (29/33) of aSAH patients without DCI and 94% (16/17) of aSAH patients with DCI were correctly assigned. Using a 99% CI and a 95% CI, 28% and 22%, respectively, were not defined, and no false-negative patients were found (Figures 1E-F). We validated our algorithm by also predicting the left-out patient samples of the 8 training sets described above. Only 6 out of the 48 excluded patients were classified in the wrong groups: 5 DCI+ and 1 DCI-. Of these 6 patients, three DCI+ patients were not determined using 95% CI, and 2 DCI+ patients were correctly classified when considering DCI score calculated on all patients. In addition, a retrospective power calculation indicated that 100% confidence at FDR of 0.05 can be achieved with a sample size of 17 patients per group (Figure S2).

Finally, the predictive algorithm was challenged against control patients who had been admitted to the NCCU but were diagnosed with other neurological disorders (Table S1). Our model correctly classified all but one of these patients as DCI-, as expected (Figure 2).

**Figure 2.**
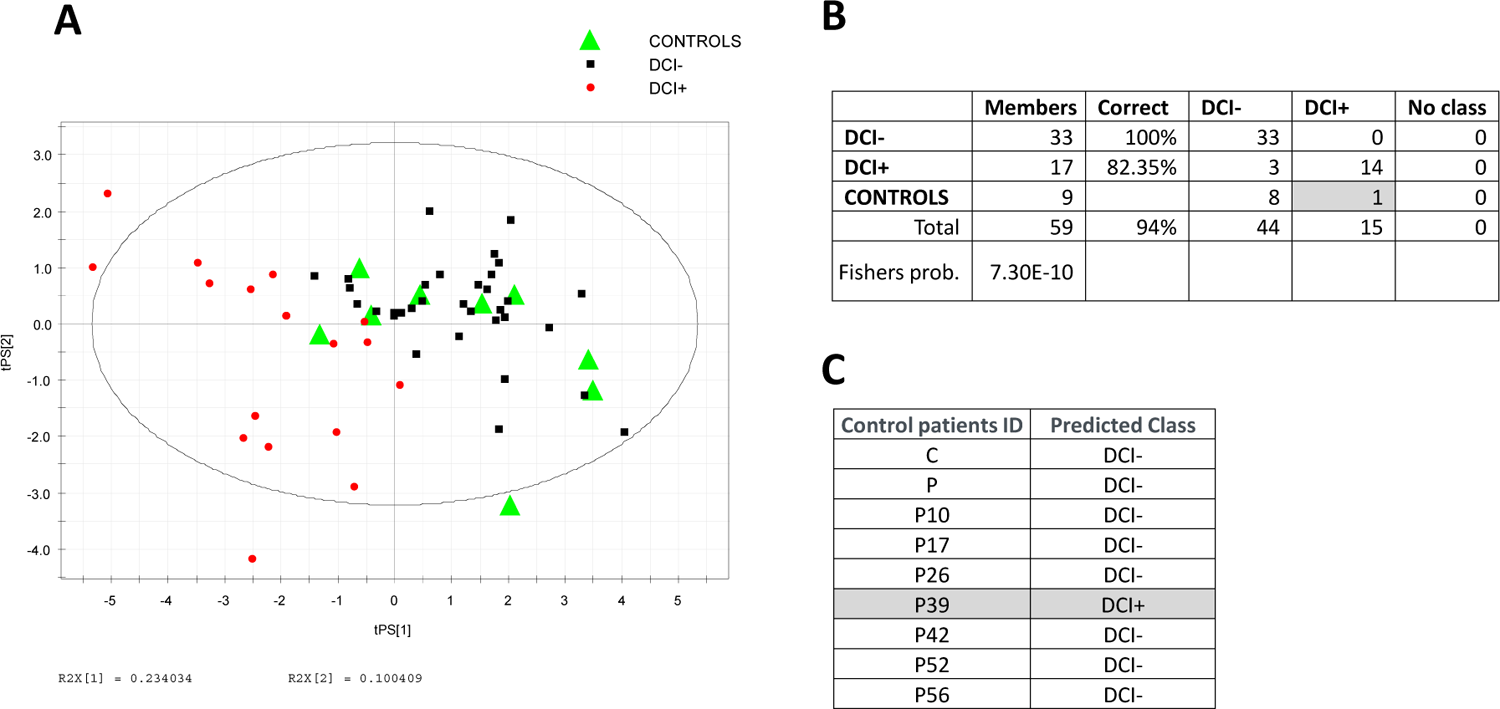
Predictive performance of the multiplex biomarker on control patients with no diagnosed aSAH. Panel **A**, DCI+ (red dots) and DCI-(black squares) are used as a PLS-DA training model to visualise control patients with no aSAH and no DCI using the multiplex biomarker. Panel **B**, confusion matrix and class assignment probability calculated by the NIPALS algorithm from the PLS-DA model in panel A. Panel **C**, class assignment of control patients calculated by the ROC.

### Multiblock analysis

We performed a multiblock analysis on all the data from the entire cohort (n=61). Annotated metabolites detected in CSF and plasma were clustered into 47 and 81 functional biological blocks, respectively, as described in the method section (5,16) (Tables S3 and S4). Lipids were blocked according to statistical proximity using hierarchical clustering analysis into 14 and 15 different blocks for CSF and plasma, respectively (5,16) (Figure S3 and supplemental Tables S5-6).

Each functional block was then analysed using a hierarchical PLS-based approach (hierarchical PLS). We tested the effectiveness of the blocking procedure to ensure it did not distort the mapping of observations in the PLS space, by comparing the PLS-DA score plots of the weighted block against the original unblocked data (5,16) (Figure S4).

Using a statistical multi-test procedure, we found a set of 14 biological functions and 8 lipid clusters in both plasma and CSF that were differentially regulated between DCI+ and DCI-patients (Figure S5).

We then set out to hierarchise metabolic functions and lipid clusters that had most impact in predicting the DCI phenotype. For that purpose, we first arbitrarily used the *t*-test *p*-values of the 14 biological functions and 8 lipid clusters previously selected (Figure 3).

**Figure 3.**
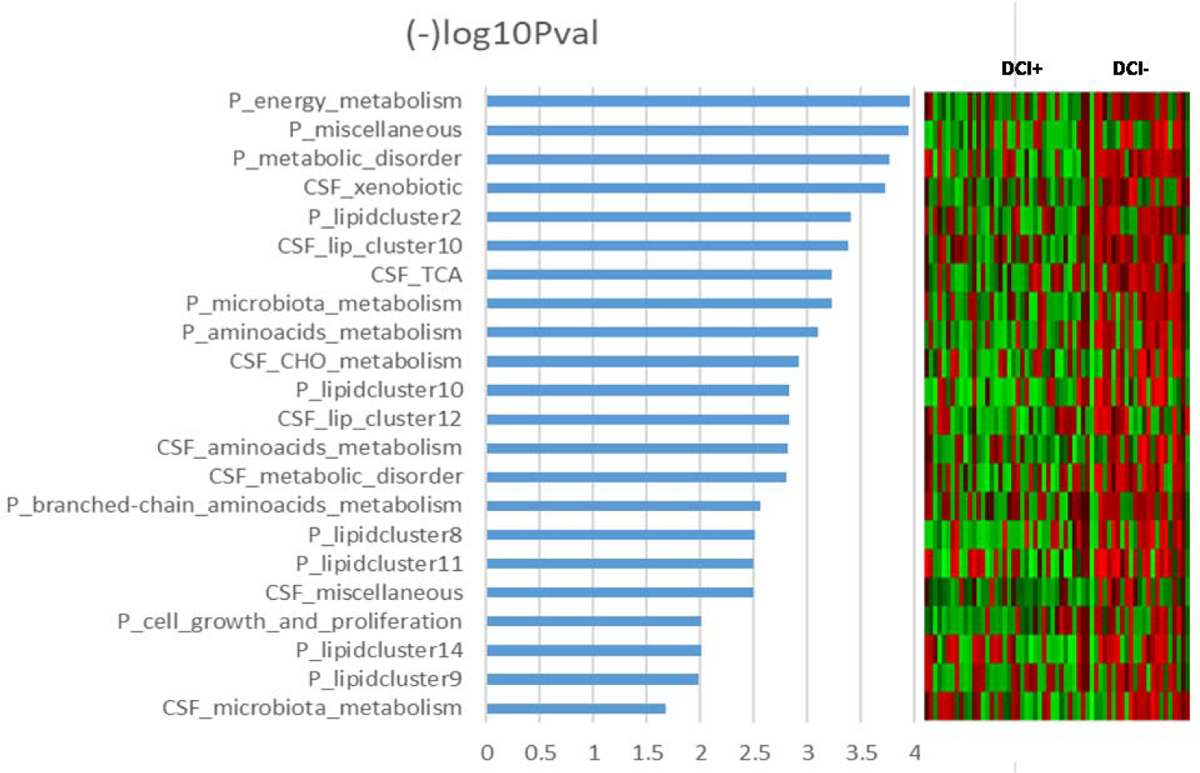
Influence of each metabolic function and lipid cluster in determining DCI status from analyses performed at t0 (first 24h post-aSAH). The variables are sorted according to their *p*-value after –log10 transformation (where low *p*-values indicating high significance correspond to high log-transformed values). Right inset, the heatmap score value of the corresponding functions and clusters for each patient. See Tables S3-6 for full details on metabolic function and lipid cluster compositions.

Given that the metabolic regulations rarely occurred independently (17), we calculated a pairwise partial correlation network integrating all the functions and clusters (Figure 4). This allowed to highlight nodes with important connectivity by calculating the coefficient of betweenness centrality, which represents the degree to which nodes stand between each other. A node with higher betweenness centrality would have more control over the network, and thus over the metabolic system. Thus, besides the statistical significance, this dimension of the network topology also affords a way to gauge how far specific metabolic regulations influence a biological system (18).

**Figure 4.**
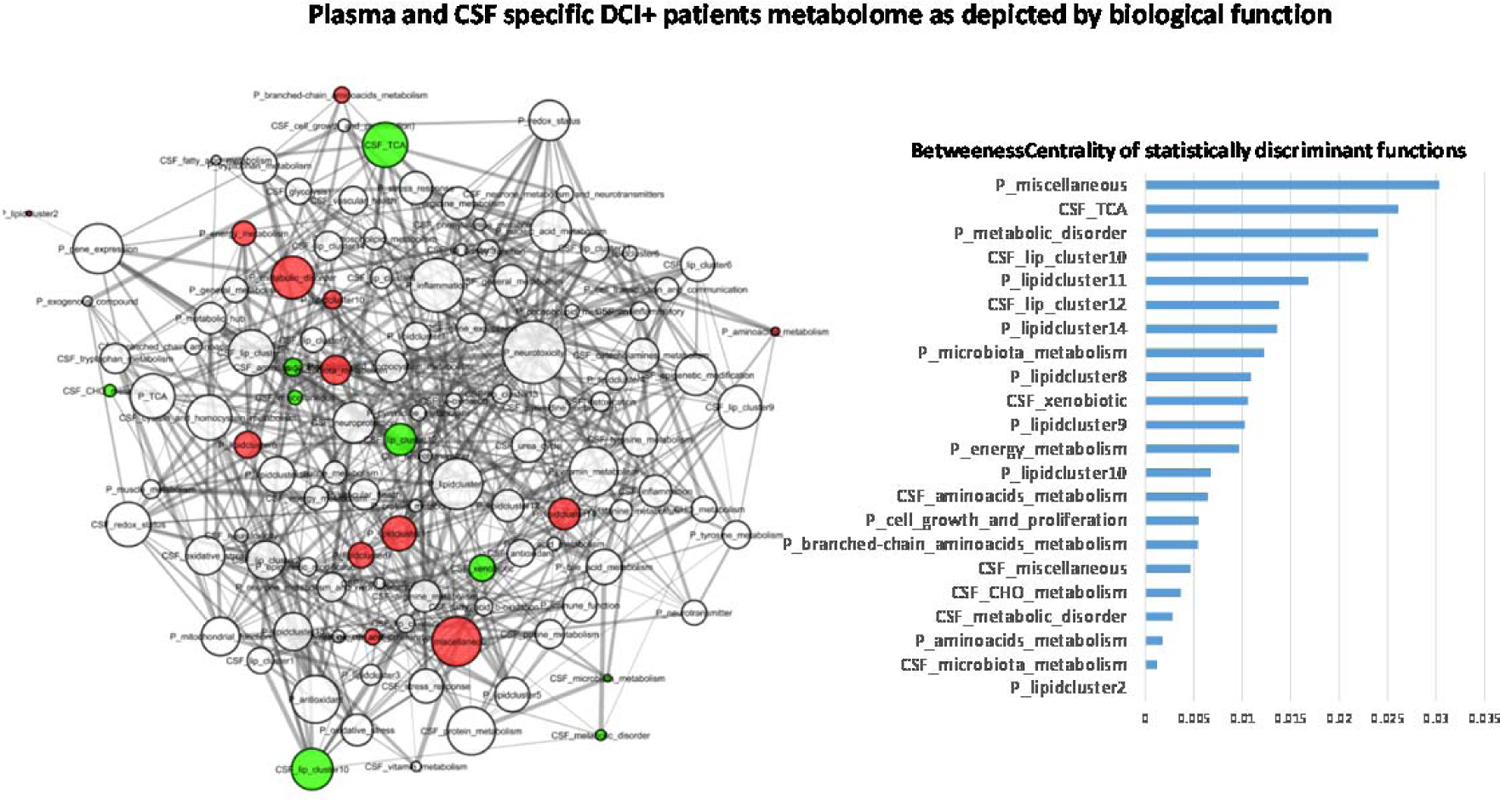
Minimum pairwise partial correlation network integrating all the metabolic functions and lipid clusters in DCI+ patients, and cleaned up from the edges of DCI-patients. The significant functions and clusters of Figure 4 are mapped in the network, with red nodes for plasma and green nodes for cerebrospinal fluid. Edge size is proportional to the *p*-value of the pairwise relationships. Node size relates to the value of the betweenness centrality coefficients calculated in Cytoscape. These coefficients used for selection of the 22 significant nodes are also reported in decreasing order in the right insight.

Finally, we merged both information by plotting for each cluster and function the betweenness centrality coefficient and the *p-*value. This allows us to highlight the regulations that most influence the metabolic systems related to the development of DCI (Figure 5).

**Figure 5.**
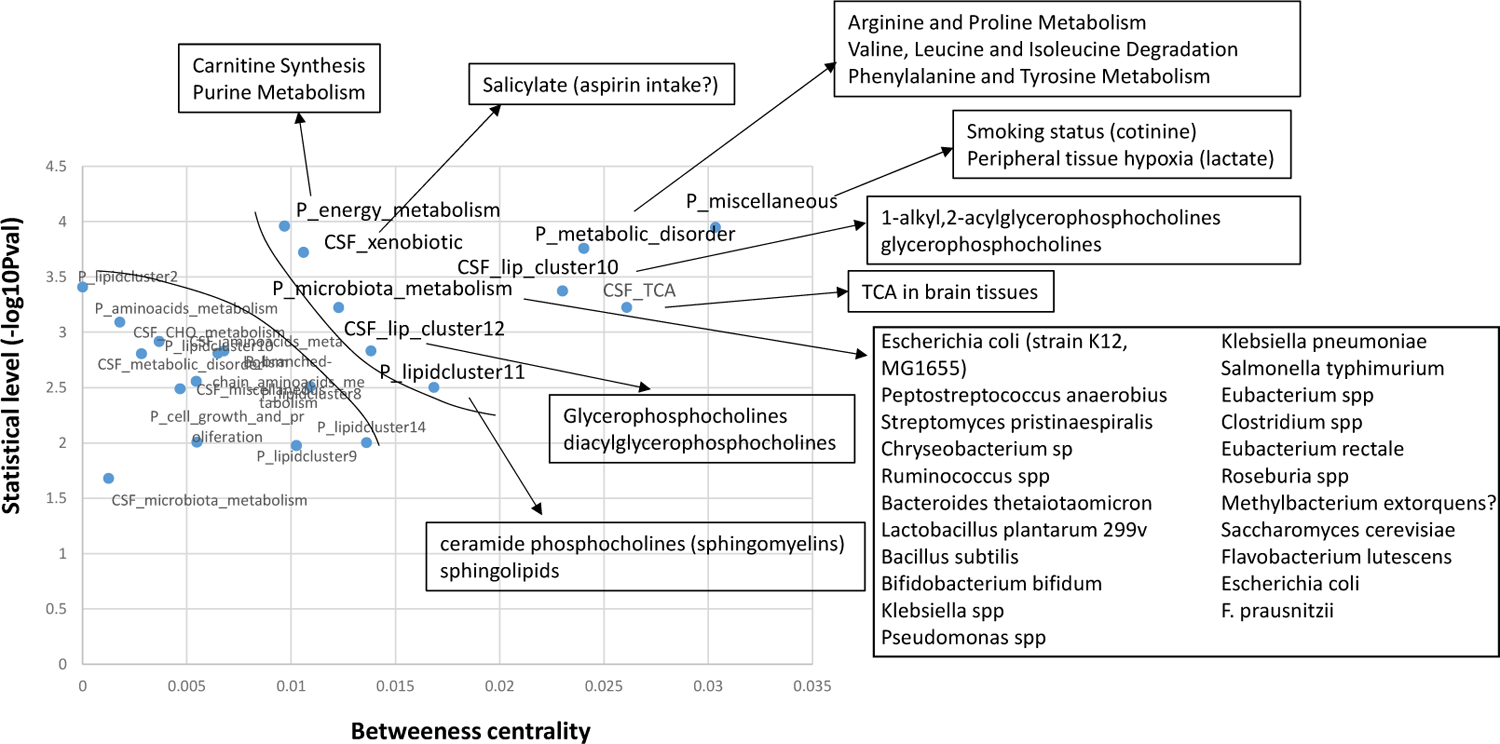
Betweenness centrality plotted against statistical *t*-test *p-value* for the 22 selected metabolic functions and lipid clusters. The most relevant variables for both P-value and betweenness centrality coefficient are highlighted in the upper-right region of the plot. The individual components of each function and cluster are analyzed using the enrichment tool where appropriate, or else in relation to their main individual variable driver (CSF_xenobiotics and P_miscellaneous) when enrichment was not appropriate. The putative bacteria producing metabolites of the P_microbiota metabolism function are reported. Links between metabolites and gut bacteria are indicated in Table S7.

Based on this dual filtering analysis (p-value and betweenness centrality), the miscellaneous score value appeared to have greater importance for determining patient DCI status. The miscellaneous cluster gathers all the metabolites left that cannot constitute functional sets (at least 3 metabolites per set). The miscellaneous score was mainly driven by both higher plasma cotinine and higher plasma lactate in DCI+ patients (Figure 5 and Figure S6).

The metabolic disorder function comprising 32 metabolites was associated with a dysregulation of important metabolic pathways such as arginine and proline metabolism, branched-chain amino-acids availability, and tyrosine and phenylalanine metabolism. Energy metabolism (comprising 8 metabolites) also emerged as important, and was related to carnitine synthesis and purine metabolism. Also, part of sphingolipid metabolism appeared to be associated to further DCI occurrence in aSAH patients.

We also found that microbiota metabolism score calculated from 18 plasma metabolites differed according to DCI status. These metabolites can be produced by 23 different bacterial species, whose metabolism possibly varies according to patient DCI status. We also found functions in CSF that were relevantly associated to the further clinical outcome but that were different to those found in plasma. Tricarboxylic acid cycle regulation appeared different between the two patient outcomes, as did two lipid clusters related to ether lipids and glycerophosphocholines lipid species. Note that a cluster comprising xenobiotics in CSF scored differently according to patient DCI status, and was mainly driven by higher CSF salicylate in DCI+ patients.

## DISCUSSION

DCI is one of the worst complications of aSAH, but also one of the most common: it occurs in approximately 30% of patients, usually between post-bleed day 5 and 14, and is associated with poor outcomes (19). DCI has a highly complex set of underlying mechanisms that include cerebral vascular dysfunction, microthrombosis, cortical spreading depolarisation, and neuroinflammation (3).

Identifying early predictive biomarkers of DCI at hospital admission would mark a huge step forward for managing aSAH patients in neurological critical care. A deeper understanding of the biological dysregulations associated DCI could lead to novel therapeutic strategies.

To address these challenges, this study had two main goals: i) to identify a shortlist of predictive biomarkers of DCI in patients admitted to the NCCU with spontaneous aSAH in blood and CSF sampled in the first 24h post-aSAH; ii) to identify early metabolic deregulations that lead to this life-threatening complication. To our knowledge, this study is the first to report the results of metabolomics and lipidomics approaches on both plasma and CSF in patients with aSAH.

We identified a set of 20 metabolites from both plasma and CSF that are putative predictive biomarkers of DCI. These metabolites were combined into an equation that generated a per-patient score that was sufficiently sensitive and selective (0.94 and 0.88, respectively, at 100% power from 17 patients per group) (Figure 1). This kind of strategy has proven valuable in other studies for defining a biomarker that is less affected by inter-individual variation or uncontrolled environmental influences (16). The multiplex biomarker score at the 99% CI was not associated with false-negative results and can therefore identify patients who are not at risk for developing DCI (Figure 1). Of interest is that independent control patients with neither aSAH nor DCI were quite confidently assigned as no-DCI cases, as expected. This substantially consolidates the relevance of our metabolite panel and score for improving DCI prognosis. Furthermore, our new multiplex biomarker shows better performance than the sole MMP-9 biomarker we previously used (15). This important key step raises prospects for replicating our findings in a multicentric study and for precisely quantifying each selected metabolite to generalise the results.

This panel of 20 metabolites is composed of 14 lipids and 6 polar metabolites. Here we found that blood lactate has the greatest discriminant power in the panel of metabolites. Nevertheless, other studies have yielded inconsistent results for blood lactate: like here, some showed that blood lactate correlates to aSAH outcomes (12,20,21), whereas others did not (22). This highlights the risk of using a single biomarker approach to predict DCI, as it can be influenced by orthogonal events.

Cigarette smoking is one of the biggest risk factors for aSAH (23–26) and for recurrent aSAH after aneurysm repair (24), and it has also been associated with symptomatic vasospasm after aSAH (27,28). Our results are consistent with these findings, as vasospasm is one of the factors contributing to DCI pathophysiology, and 90.1% of the DCI+ patients in our study experienced a vasospasm. Cotinine is a nicotine metabolite that is widely used as a biomarker of cigarette smoking. Here we found a higher level of cotinine in DCI+ patients, whereas the reported smoking status by interview appeared less obvious (Table S2). This could be due to underreporting by patients or to passive smoking.

The reduced set of the 20 metabolites identified through this biomarker approach remains insufficient to provide a mechanistic explanation for complex disease phenotypes. Therefore, in order to push forward and decipher the metabolic dysregulations associated with DCI, we employed multiblock analysis and enrichment analysis that together make it possible to summarise complex metabolite profiles into meaningful functions or pathways as well as to plot the relationships of each set of functions and clusters in an interaction network. Then, crossing the statistical impact of the functions/clusters with the interaction network topology helped to hierarchise the impact of each biological module on DCI occurrence (Figure 3). For instance, the “miscellaneous” cluster of metabolites had the best statistical power associated to the influence of metabolic system (betweenness centrality) for defining DCI status. Since this cluster was mainly driven by plasma cotinine and plasma lactate, it again highlights the major influence of early peripheral tissue hypoxia together with smoking status for predicting a poor-prognosis DCI event, as observed elsewhere (12). Note that the xenobiotics cluster in CSF was significantly predictive of DCI status, and much of this predictive power was driven by higher CSF salicylate in DCI+ patients. Salicylate likely arose from aspirin intake and its subsequent deacetylation (29), which would be higher in DCI+ patients and may well represent a surrogate marker of frequency of headache at aSAH onset. It could be important to monitor aspirin intake and the underlying reasons for aspirin use in aSAH patients.

Enrichment analysis of lipid species in specific relevant lipid clusters pointed to a lower level of some subspecies of sphingomyelins in plasma and a higher level of some phosphatidylcholine subspecies and derivatives in both plasma and CSF in DCI+ patients (Figures S7-8). It is not possible at this stage to establish whether a metabolic deregulation of a specific lipid directly drives DCI or is only a surrogate mechanism. However, as sphingomyelins are metabolised in ceramides (CER) by sphingomyelinase (SMase), a lower level of sphingomyelins could be a consequence of a higher SMase activity. Testai et al. (8) found an elevated level of ceramides, particularly C18:0, within 48 hours post-bleed in the CSF of patients with symptomatic vasospasm. CER are important mediators of apoptosis, and several studies have described increased CER levels in stroke patients (30). Moreover, SMase-driven CER production induces IL-6 expression (8,31) and triggers the vasoconstrictive properties of sphingolipids (32,33). Further studies are needed to determine whether lower plasma levels of specific sphingomyelins in DCI+ patients are linked to enhanced ceramide production and occurrence of vasospasm.

The metabolites in three biological functions, namely metabolic disorders and energy metabolism in plasma, and tricarboxylic acid cycle (TCA) in CSF, can be translated into metabolic pathways using enrichment analysis. These functions related specifically to arginine and proline metabolism, phenylalanine and tyrosine metabolism, branched-chain amino-acid degradation, carnitine synthesis, and purine metabolism. Greater TCA cycle deregulation in the brain might well be related to the hypoxia described by high plasma lactate levels, reflecting a greater global impairment of aerobic respiration, with mitochondria as the main target (34). This global impairment might be also related to the difference in energy metabolism attached to carnitine and purine metabolism and measured in plasma between the future DCI+ and DCI-cases. Other impairments also applied to the metabolic deregulations observed in plasma that were related to arginine and proline, phenylalanine and tyrosine, and branched-chain amino-acid metabolisms. NO bioavailability is central to controlling brain perfusion in DCI (35) and is related to arginine metabolism.

Phenylalanine and tyrosine metabolism is related to catecholamine synthesis, and high plasma catecholamine levels were found to be related to poor aSAH outcomes (35). Our observation further highlights the relationships between aSAH catecholamine mediated stress with high circulating lactate hypothesised by Van Donkelaar et al. (12). Note that rat studies have found a relationship between aberrant branched-chain amino acid metabolism and ischemic stroke. This relationship was due to a dysregulation of the microbiota–gut– brain axis (36). No such mechanism associating BCAA and microbiota has yet been reported for aSAH and DCI, but our finding that BCAA metabolism is linked to microbiota metabolism potentially involving bacterial species is consistent with (36). Most previous studies on aSAH and DCI have focused on brain metabolic issues. Most of them, with the exception of (12), have largely ignored the relationship of brain metabolic impairments with peripheral metabolism. It is established that such relationship greatly influences the incidence and severity of ischemic stroke and can impact post-ischemic stroke outcome (37). Our results in aSAH patients also pointed such interaction and integration between peripheral and cerebral metabolism, as touched on elsewhere (12). How this peripheral impairment would affect DCI occurrence remains unknown. It could also be a reflection in plasma of metabolic deregulation in the brain following disruption of the blood–brain barrier (34), and could be specific to poor DCI outcome.

The advantage of our approach is that it is possible to hierarchise the importance of each of the above-discussed events and metabolic deregulations associated to DCI outcome, as illustrated in Figure 4. This hierarchisation confirms that smoking status and peripheral hypoxia are the main risk factors, followed by peripheral metabolic deregulations likely associated to NO, catecholamine and BCAA metabolism, but also impairment of the tricarboxylic acid cycle in cerebral mitochondria and some phosphorylcholine lipids. Finally, other factors also emerged as slightly less important determinants of DCI, such as some aspects of peripheral sphingolipids metabolism and energy metabolism, gut microbes, and possibly also aspirin intake.

In conclusion, we identified a high-performance predictive metabolomic/lipidomic multiplex biomarker of future DCI in aSAH patients at admission into neurological critical care. The integrative approach adopted here also highlighted important biological (both peripheral and cerebral) deregulations associated to DCI occurrence, including peripheral hypoxia (blood lactate), specific lipid metabolism alterations, and deregulation of important metabolic functions. Analysis also pointed to possible exogenous causes of DCI, such as smoking and aspirin intake. Furthermore, our results importantly point for the first time to a possible gut microbiota–brain axis of DCI. The exploratory research reported here is based on a single-centre study. This important first step lays the foundations for a multi-centric study to validate our results in different populations.

## Data Availability

All the data used in the manuscript are available upon request to, except those excluded for ethical reasons.

## Acknowledgements

We thank the staff at the NCCU and Marseille La Timone hospital laboratory for managing the sampling.

## List of supplemental materials

### Supplemental methods

Tables S1-S7

Figures S1-S8

References: 38-41

## Non-standard abbreviations and acronyms

WFNS: World Federation of Neurological Surgeons

IVH: intraventricular haemorrhage

PCA: principal component analysis

PLS-DA: partial least squares–discriminant analysis

GOSE: Glasgow Outcome Scale Extended

VIP: variable importance in projection coefficient

FDR: false discovery rate

HILIC: hydrophilic interaction liquid chromatography

RP: reverse-phase chromatography

